# Mosquito Species Composition and the Risk of Vector-Borne Disease at Live and Wet Markets in Laos

**DOI:** 10.1101/2025.03.24.25324509

**Authors:** Elin Asp, Patrick Höller, Julia Pärssinen, Nalita Adsanycnah, Khamsing Vongphayloth, Somsanith Chonephetsarath, Vannaphone Phuttana, Mahmoud M. Naguib, Johanna F Lindahl, Jiaxin Ling

## Abstract

Live and wet markets (LWMs) form an important pillar of food supply in Asian countries, but the close interaction of animals and humans at markets creates an interface for disease transmission. The zoonotic risks associated with LWMs were highlighted after the onset of the COVID-19 pandemic. This study aimed to investigate the risk for mosquito-borne disease transmission at LWMs, through analyzing mosquito species composition and prevalence of flaviviruses, as well as general perception of the mosquito-borne diseases at LWMs in Lao PDR. Adult mosquito- and larval samples were collected from 15 LWMs. A total of 1129 adult mosquitoes were collected from five genera, of which *Culex* was the most common genus, with *Cx. quinquefasciatus* (*qq*.) being the most abundant species (85%). Moreover, 188 *Culex* larvae/pupae were collected, with a majority ovipositioned in water accumulated by the activities of LWMs, such as frequent water and ice distribution to keep the food fresh. All collected mosquitoes were grouped into 184 pools and were analyzed by a Pan-Flavi RT-PCR assay, with sequencing of suspected positives. No pathogenic flaviviruses were found from collected mosquitoes; however, mosquito-specific orthomyxoviruses were identified. The study questionnaire indicated a high awareness regarding mosquitoes as a source of disease transmission. Conclusively, accumulated water from LWMs is a focal point for mosquito oviposition and propagation, especially in larger-sized markets, enabling the zoonotic transmission of mosquito-borne diseases. This suggests implementing mosquito-mitigation measures such as wastewater management to prevent diseases associated with mosquitoes.

**Author Summary:** Mosquito-borne diseases, especially flaviviruses, have had a rapid increase in Asian regions in the past decade. Live and wet markets (LWMs) are common sources for food in Asia and have repeatedly been associated with the risk of zoonotic transmission, especially after the COVID-19 pandemic. We aimed to identify the mosquito species and explore associated viruses circulating the markets in Lao PDR, to evaluate the risks of mosquito-borne diseases. Among collected mosquitoes, *Culex* was recorded as the most prevalent genera. The most common mosquito oviposition site was stagnated water in the market, especially in larger LWMs. One larval pool was found positive to two mosquito-specific orthomyxoviruses, but no pathogenic flaviviruses were detected. This study demonstrates that live and wet markets a common site for mosquito circulation and oviposition, highlighting the risk of mosquito-borne disease transmission at such markets.

## Introduction

Mosquito-borne flaviviruses cause many important zoonotic diseases globally, and include dengue virus (DENV), Japanese encephalitis virus (JEV), Zika virus (ZIKV), West Nile virus (WNV), and yellow fever virus (YFV) (1). These single-stranded RNA viruses have had a rapid emergence in the past decades of which more than half the world’s population is at risk of infection (1,2). Asia is particularly affected by mosquito-borne flaviviruses, accounting for 70% of the global dengue burden, with a 46% increase in DENV cases from 2015 to 2019 in Asia (3). Additionally, Asia serves as the primary area of JEV transmission (4). The globally increased flavivirus transmission is driven by factors such as climate anomalies with increased temperatures, urbanization, and increased traveling (5). With inadequate treatment options and increased flavivirus transmission, despite vector control measures, understanding the risks and mitigation options becomes even more important (5).

Flavivirus infections commonly present as febrile illness, with risk of more dangerous clinical manifestations such as hemorrhagic fever and encephalitis (1). Despite similar physiopathological patterns, the flaviviruses have different transmission routes and host ranges. Dengue and Zika are transmitted between humans by *Aedes* mosquitoes, which is also why highly populated cities tend to be endemic to the disease (5,6). In the transmission cycle of *Aedes* mosquitoes, oviposition commonly occurs in artificial water containers and the mosquito resides mainly indoors (7). JEV, on the other hand, is transmitted by the nocturnal *Culex* mosquitoes which prefer stagnant or fresh water for oviposition (5,8). JEV is transmitted in an enzootic transmission cycle between waterfowl as the natural host and pigs as the amplifying host (4), and humans are occasionally subjected to zoonotic transmission and thus, act as a dead-end host (4).

Live and wet markets (LWMs), highly prevalent in Asian countries, are a source of local trading of food and live- or slaughtered animals (9). Thus, these markets hold socio-economic importance in Asia (9) while also facilitating an interchange between live animals, humans, and the environment (10,11). LWMs have been in the spotlight due to their risk of disease emergence after the COVID-19 pandemic (2019–2022) (9). The zoonotic risk is driven by the close interaction between animals of wild and domestic origin with vendors and visiting customers at the markets (9). Vendors keep produce and fish fresh by frequent water and ice distribution, risking accumulation and stagnation of the water when draining systems are inadequate. Water accumulations from the markets have the potential of creating breeding grounds for mosquito oviposition (9,11). With LWMs commonly situated in urban foci, there are many potential hosts for pathogenic viruses, causing a risk of disease transmission. Thus, these factors can increase the risk of transmission of mosquito-borne diseases at LWMs, facilitating both urban transmission of DENV and ZIKV, as well as enzootic and epizootic transmission of, e.g. JEV (9,12).

Lao PDR (People’s Democratic Republic), a landlocked country in Southeast Asia, has approximately 400 towns with LWMs with frequent trade of international and domestic wildlife (13). This puts Lao PDR in a vulnerable state with a risk of zoonotic transmission (13,14). Moreover, Lao PDR is hyperendemic of all four DENV serotypes (DENV 1–4), with the capital city Vientiane reporting the highest burden of dengue in the country (15,16). The country has additionally experienced an upsurge in DENV cases over the years, with 35 000 reported cases in 2023, an increase of 5000 cases compared to the previous year (17,18). The risks of mosquito-borne viruses at LWMs in Lao PDR, or any Asian country, have yet to be extensively investigated. Thus, this study aims to study the perception and distribution of mosquitoes at LWMs in the district of Vientiane, in Lao PDR, and screen the mosquitoes for flaviviruses.

## Methods

### Ethical Considerations

The project received the ethical permit from the national ethics committee of Lao PDR (number 032/NECHR). The provincial or district health staff or market directors approved the markets’ sample sites. Prior to installation of the mosquito traps, respective vendors were informed about the study and gave verbal consent.

### Study Design

LWMs, for sample collection, were in agreement with staff from the provincial markets and the National University of Laos (NUOL). The markets were categorized based on the number of vendors in the market. small-sized market: 1–14 vendors; medium-sized market: 15–100 vendors; large-sized market: > 100 vendors. The entomological collection was performed during the dry season from February 1^st^ – March 9^th^, 2023 at LWMs throughout Vientiane and Vientiane province, Lao PDR. A total of 15 markets were subjected to sample collection, of which 4 were small-sized markets, 5 were medium-sized markets, and 6 were large-sized (Fig 1A, B). Of note, some markets yielded only adult mosquitoes, others only larvae, and some both. The species distribution of live animals kept and sold at the LWMs was recorded (Fig 1C).

**Fig 1.**
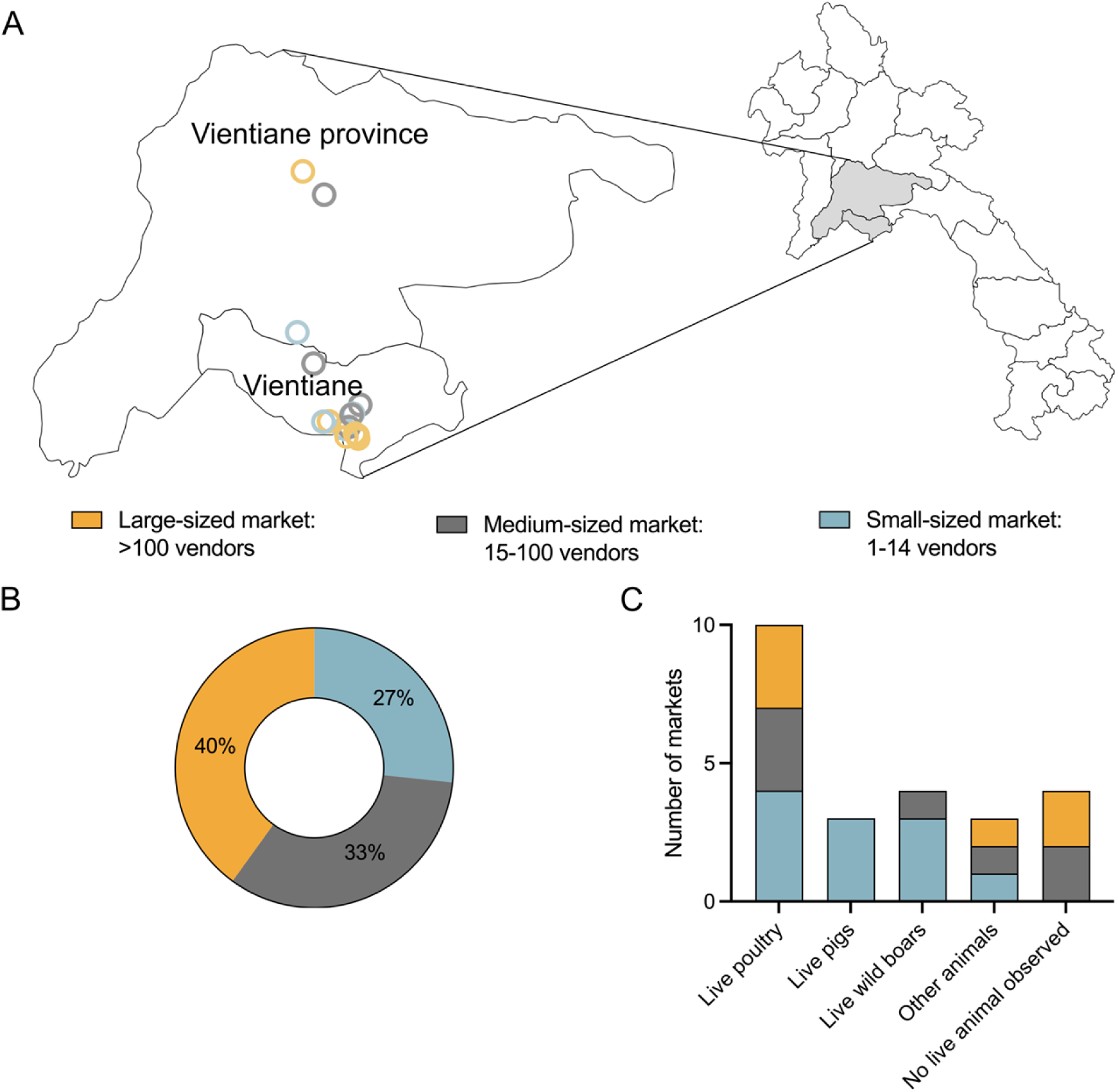
Overview of the sampling at LWMs. (**A**) Sampling locations according to market size in Vientiane capital and Vientiane province, Lao PDR. (**B**) Distribution of the differently sized LWMs. Large-sized markets in yellow, medium-sized markets in gray, and small-sized markets in blue. (**C**) Live animals observed at the LWMs and at what market size. Other animals are squirrels, rabbits, goats, pigeons, and birds of unknown species.

In addition to the entomological collection, a questionnaire survey was conducted to assess the knowledge of mosquito-borne disease and perception of mosquito-biting frequency at the LWMs. The survey included 52 vendors from 12 LWMs, of which five of these LWMs were not included in the entomological collection. Vendors were randomly selected for the interview by the local veterinary staff of NUOL using the local language.

### Mosquito and Larval Collection

Mosquitoes were collected using CDC (Centers for Disease Control and Prevention; John W. Hock, Gainesville, Florida, USA) light traps and a hand net. Due to the limited local resources, CO_2_ bait was not used with the CDC light trap. The CDC light traps were placed at the LWMs in the morning from 6–11 am and collected the following day, after 24 ± 4 hours. The hand net was used throughout the market, focusing mainly on corners and vendor stalls, for 15 minutes at any point during the day. Collected mosquitoes were stored at -20 °C and transferred to falcon tubes following euthanasia.

Different water bodies at the LWM were examined for mosquito larvae and pupae. Following the inspection, one or more mosquito larvae were collected from the water bodies using a plastic dipper. Collected larvae were placed in 70% ethanol in 1.5 ml tubes and stored in a cooling box for a maximum of six hours, awaiting storage at -20 °C. Mosquito and larval samples were labeled with collection site, sample ID, and date.

### Species Identification and Pooling

The identification was conducted by entomologists at the Institute Pasteur du Laos. Adult mosquitoes were morphologically identified at species level and mosquito larvae and pupae were identified at genus level using available keys (19–21). The specimens were pooled based on sample site, species, and sex, with a maximum of ten specimens per pool. The blood-fed female mosquitoes were separated, creating individual pools. The samples were homogenized using sterile glass beads or metal beads, with a TissueLyser. 400 µL media was used for each tube, composed of phosphate-buffered saline (PBS; 137 nm/L NaCl, 2.7 nmol/L KCl,10 nmol Na_2_HPO_4_, 1.8 nmol/L KH_2_PO_4_) supplemented with 100 mg/L gentamycin, 100 mg/L streptomycin, and 100.000 U/L penicillin G. The samples were homogenized at 20 000 rpm for three minutes followed with centrifugation at 9000 rpm for three minutes. A total of 184 pools were created and preserved at -20 °C awaiting further processing.

### RNA Extraction and Pan-Flavi Assay

RNA from the samples was extracted from 140 µL homogenate using the QIAamp®Viral RNA Mini Kit (Qiagen®, Hilden, Germany) according to the manufacturer’s instructions. The samples were stored at -80℃ until further analysis. The extracted RNA was converted to cDNA using the high-capacity cDNA reverse transcription kit (Applied Biosystems). Flavivirus screening was performed using a pan-flavi assay with three universal primers (S1 Table) (10 pM), as previously described by Patel et al. (22). Samples were run on a qTOWER3 thermal cycler (Analytik Jena) with qPCRSoft3.4 software at NUOL and then confirmed on the CFX96 Real-Time System (BioRad), at Uppsala University.

### Gel Electrophoresis and Sequencing

PCR products that were suspected positive in the RT-qPCR with CT values below 40 were run on gel electrophoresis. PCR products of expected amplicon size (266 base pairs) were sent for Sanger sequencing to Macrogen Europe (macrogen-europe.com), alongside the primers. To further verify if JEV is present in those suspected positive samples, we performed a qPCR based on specific primers targeting at least JEV genotypes I and III (Forward_2514 5-GATGAGGTGTGGAAGTGGCA-3, Reverse_2770 5-GATATCTCCCCACGGGCTTG-3) (S1 Table). Sanger sequencing data was edited using BioEdit Sequence Alignment Editor to improve sequence quality. The sequences were thereafter compared with the databases of nBLAST (blast.ncbi.nlm.nih.gov/Blast.cgi). An E-value ≤ 10e^−5^ was considered significant.

### Data Analyses

GraphPad Prism version 10.0 (GraphPad Software Inc) was used for data analysis. Difference between market types and habitats was tested with t-test in STATA 15.2 (STATAcorp, College Station, Texas)

## Results

### Entomological Species Identification of Adult Mosquitoes

A total of 1129 adult mosquitoes were collected, belonging to five genera. Of these, *Culex* was the most abundant genus, accounting for 1119 mosquitoes (99.1%). Within the *Culex* genus, *Cx. quinquefasciatus* was the most abundant species of 959 specimens (84.9%), followed by *Cx. hutchinsoni* with 126 mosquitoes (11.2%), and *Cx.vishnui* with 23 mosquitoes (2.0%). The remaining species were *Cx. gelidus* (n = 5), *Cx. fuscocephala* (n = 4), *Aermigeres subalbatus* (n = 4), *Cx. sitien* (n = 2), *Ae*. *vexans* (n = 2), *Anopheles aconitus* (n = 2), *Aedes albopictus* (n =1), and *Mansonia* (n = 1) (Fig 2A). The vast majority (n = 937, 83%) of the mosquitoes were collected from large-sized markets (Fig 2B). 27 mosquitoes had blood meals and were pooled separately. Three of the six blood-fed pools originated from pig-rearing LWMs (S2 Table).

**Fig 2.**
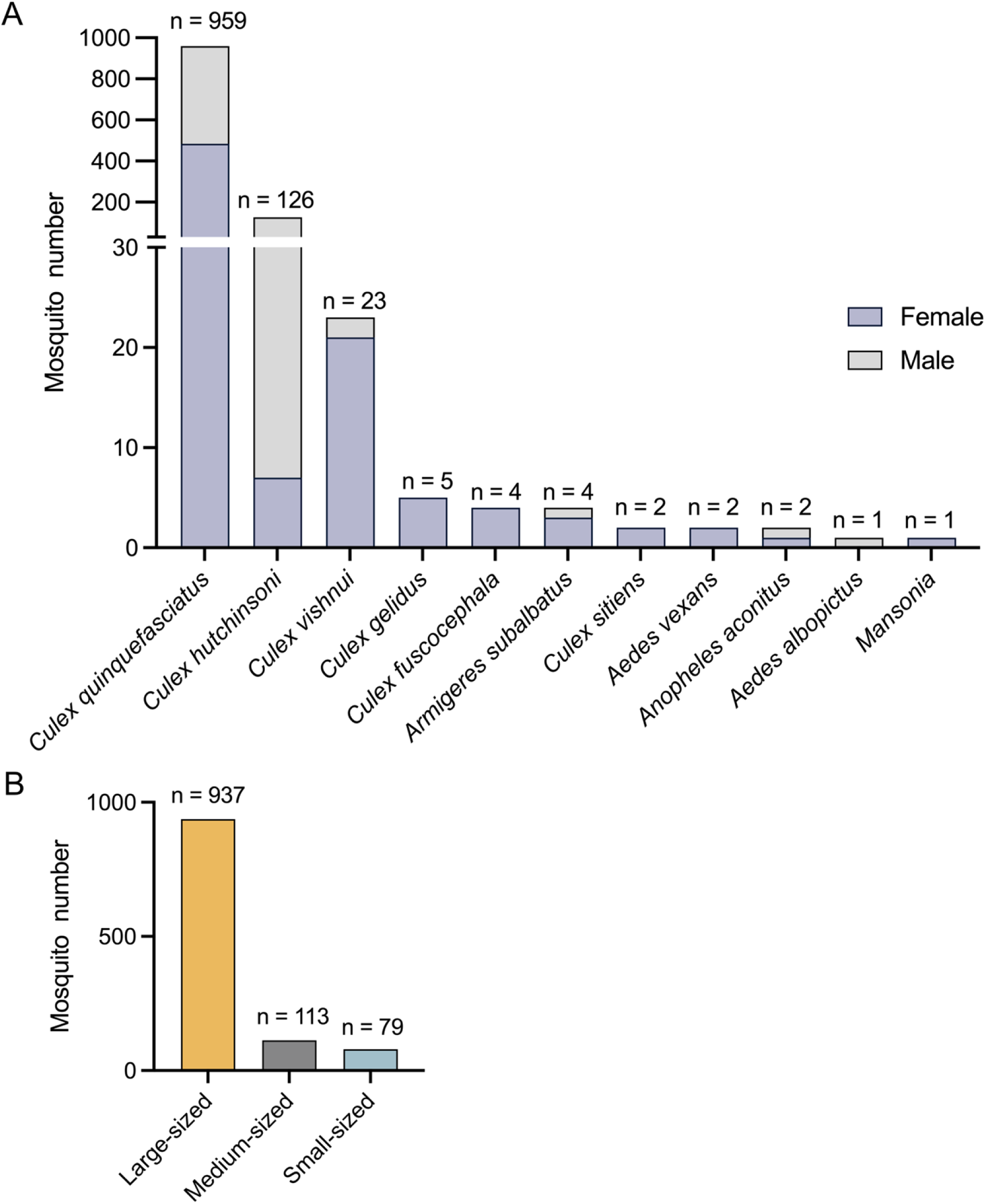
Adult mosquito collection at LWMs in Lao PDR. (**A**) Number and species of mosquitoes collected. Purple indicates female mosquitoes and grey indicates male mosquitoes. (**B**) Distribution of collected mosquitoes at differently sized markets. The number of markets used for adult mosquito collection was 6, 4, and 2 for the large, medium, and small-sized markets, respectively.

### Spatial Distribution of Mosquito Larvae and Their Habitats

Mosquito larvae were additionally collected at the LWMs. Three classes of larval habitats were distinguished: stagnant water bodies in vegetation (ponds, ditches, puddles), running water produced by the LWMs (drains), and stagnant water bodies produced by the LWMs (drains, man-made containers), seen in Fig 3A. A total of 188 specimens (larvae and pupae) were collected, all belonging to the *Culex* genus (S2 Table). Approximately half of the specimens (n = 84, 44.7%) were collected from large-sized markets (Fig 3A). The stagnated water in drains at LWMs was the most common source of mosquito breeding, accounting for almost 60% of the total larvae collected (n = 112) (Fig 3 A, B). Larvae/pupae collected in market-produced water sources were predominantly associated with larger- and medium-sized markets, in addition to having a higher specimen count per market (Fig 3B). No significant differences between market types, and not between habitats

**Fig 3.**
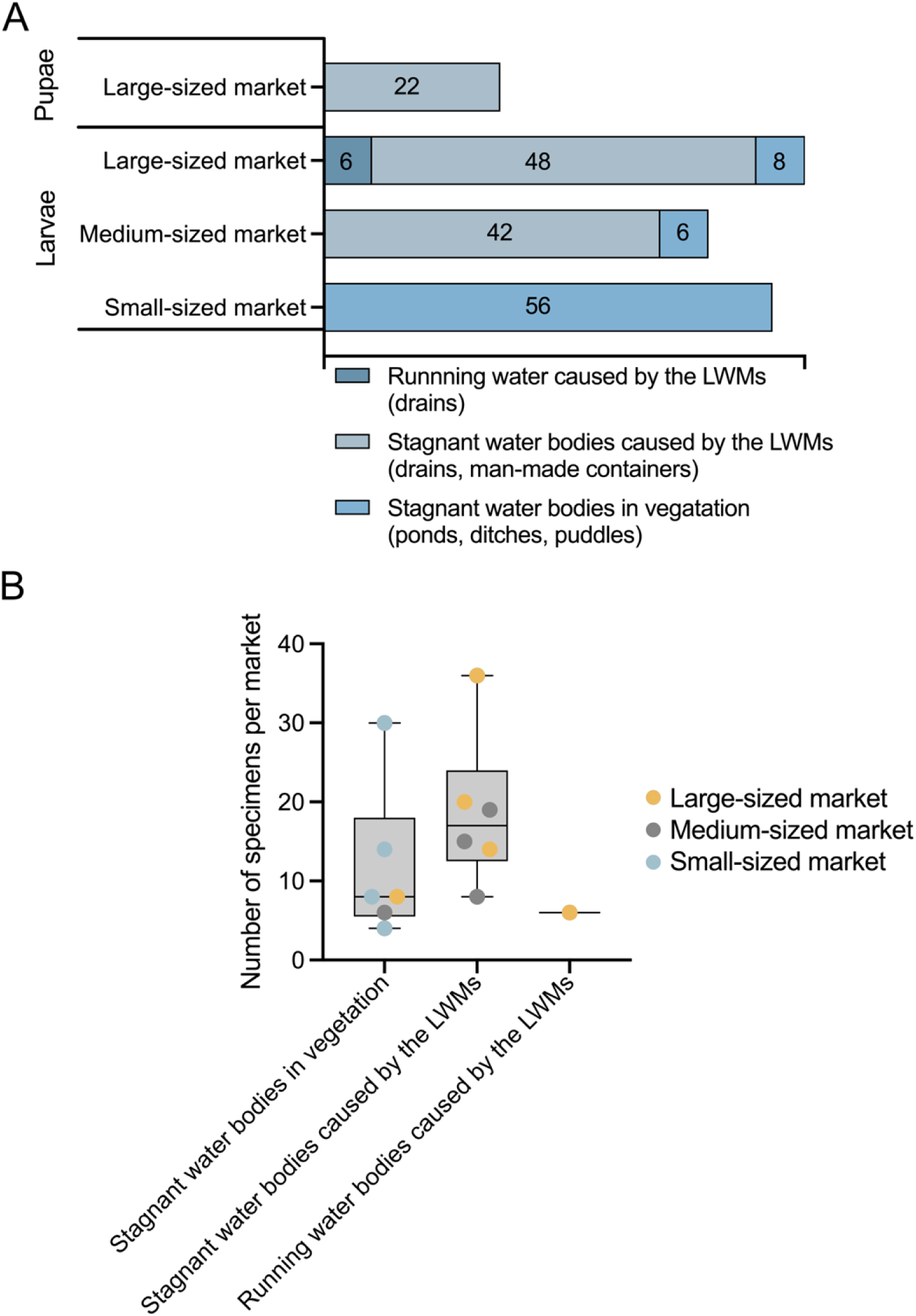
Larvae and pupae collection sites. (**A**) Overview of the number collected mosquito larvae and pupae at different market sizes and habitats. (**B**) Box plot of the number of specimens (mosquito larvae and pupae) collected from water bodies produced by the LWMs (stagnant and running water) and those collected in vegetation at LWMs.

### Screening results for flaviviruses

The 284 pools comprising 1317 specimens (S3 Table) were screened for flavivirus genus by RT-qPCR. A total of 29 pools were deemed as possible positive by RT-qPCR and were subjected to gel electrophoresis. Bands of 266 bp were detected in PCR products from 21 sample pools (S1 Fig). These 21 samples were further confirmed by using qPCR specifically targeting JEV of which none were positive. Moreover, sequences from these PCR products were obtained by Sanger sequencing. One larval pool (pool 166) showed significant similarities with two orthomyxoviruses; Wuhan Mosquito virus 6 strain at 98% identity (90 bases; e-value 2e-34) and a 93% (100 bases, e-value 2e-33) similarity with a *Cx. pipiens* orthomyxo-like virus isolate (S3 Table).

### Mosquito-Biting Frequency and Mosquito-Borne Disease Knowledge at LWMs in Lao PDR

The vast majority (90%) of the vendors were aware that mosquitoes can transmit diseases, with malaria (43%), followed by dengue fever (21%) being the only reported known mosquito-borne diseases (Fig 4 A, B). When asked about mosquito biting frequency at the LWMs, more than half of the respondents (60%) reported being bitten often, while 14% reported to get bitten sometimes, and only 7% stated to not being bitten (Fig 4C).

**Fig 4.**
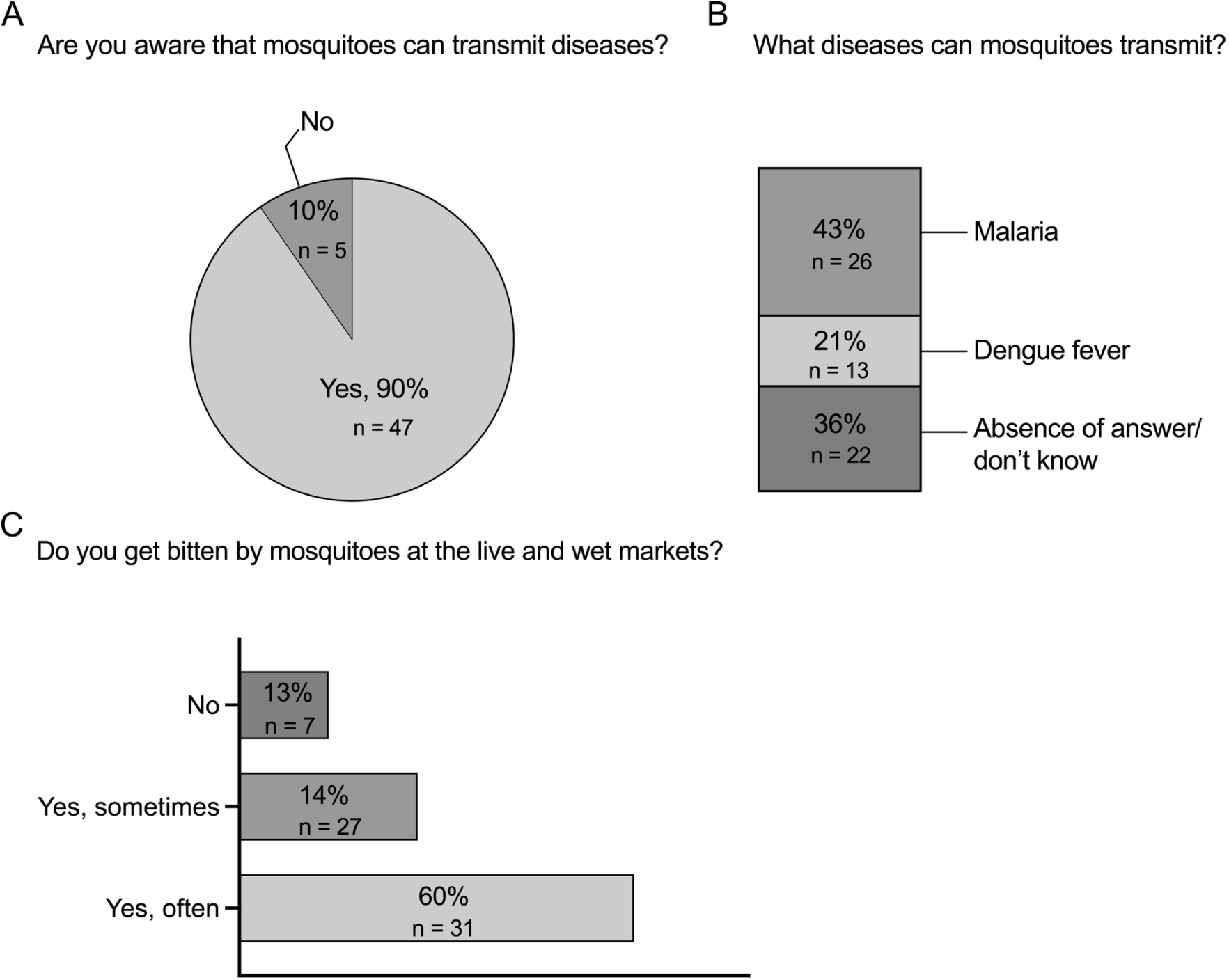
Mosquito biting frequency and awareness of mosquito-borne diseases amongst vendors in Lao PDR (**A**) Knowledge of the mosquito-borne diseases among vendors. (**B**) Proportion of vendors who identified specific mosquito-borne diseases. Some vendors mentioned more than one disease. (**C**) Mosquito biting frequency among vendors.

## Discussion

Asia has a high burden of mosquito-transmitted flaviviruses and the region’s high abundance of LWMs increase the risk of disease propagation and zoonotic spillover (2,9,10). However, the mosquito-borne disease risks at LWMs remain unclear. This study aimed to assess the role of LWMs in mosquito habitats, species, and the transmission risks of mosquito-borne flaviviruses, focusing on Lao PDR.

The high density of *Culex* mosquitoes, especially larger-sized markets, observed in this study suggests that stagnant wastewater, commonly found in the LWMs, plays a significant role in creating breeding sites for mosquitoes. This could indicate an increased risk of mosquito-borne disease transmission, especially at larger-sized markets given the high number of people frequenting the markets daily. *Cx.quinquefasciatus*, a species known to feed on both humans and animals, poses an increased risk of JEV transmission. This species can opportunistically feed on pigs, facilitating spillover of JEV to humans (23,24). Given that pigs are amplifying hosts for JEV, their presence further elevates the risk of transmission, particularly in urban LWMs (4). While *Cx.tritaeniorhynchus* is recognized as the primary vector of JEV (25), the role of *Cx.quinquefasciatus*, an abundant mosquito in our collection, as a potential vector has been emphasized, and several studies have identified JEV positive *Cx.quinquefasciatus* (25,26). Other species considered secondary vectors of JEV include *Ae. japonicus*, *Cx.fuscocephala*, and *Cx.vishnui*, among others, also present in the mosquito collection (27). Although the primary vector of JEV was not collected, the presence of other potential *Culex* vectors suggests that LWMs may still pose a risk of JEV transmission. Of importance to note, some markets were subjected to more than one adult mosquito sampling, which could create a sampling bias.

As described earlier, the *Culex* genus constituted the majority of the mosquito collection. However, *Aedes* mosquitoes, the primary vectors of DENV, would have been relevant to screen for flaviviruses, especially since DENV often spreads in urban areas (2). The large sampling bias towards *Culex* spp. (species) is most probably a result of the CDC light traps, attracting nocturnal *Culex* mosquitoes. For more effective *Aedes* sampling in future studies, BG sentinel traps or CO_2_ attractants are recommended for day-active mosquitoes like *Aedes*. Nonetheless, the larger-sized LWMs had enclosed or semi-enclosed structures, with high circulation of people, which indeed could create a favorable environment for the indoor-biting *Aedes* mosquitoes (7). To better assess the risk of flavivirus transmission at LWMs, further sampling should be conducted during the high-precipitation season, when dengue cases peak from May to October (28); also shown by a study indicating a correlation with the DENV incidence, high temperatures and moderate rainfall in Lao PDR (29). The DENV incidence is particularly high in Vientiane capital, as the largest urban area in Lao PDR, which alone had close to 40 000 reported cases in the 2019 outbreak (28). To increase the number of collected specimens and improve the species diversity, future studies should consider changing trapping method and season for entomological collection.

Moreover, the study found that the primary breeding sites for mosquitoes in LWMs were water bodies generated by market wastewater, particularly in larger markets where stagnant water was more prevalent. The presence of mosquito larvae in these water bodies suggests that poorly managed wastewater contributes to the high density of *Culex* mosquitoes. Smaller markets, in contrast, had fewer mosquitoes, with larvae primarily found in vegetation, indicating that market size and infrastructure may influence mosquito prevalence. The role of stagnant wastewater in mosquito breeding highlights the need for public health interventions, including improvements in wastewater handling to reduce mosquito habitats and mitigate the risk of mosquito-borne diseases in LWMs.

Genetically, Sanger sequencing revealed two orthomyxoviruses in one larval pool: Wuhan Mosquito Virus 6 strain and an isolate of *Culex pipiens* orthomyxo-like virus. These are non-pathogenic insect-specific viruses and have been found in mosquitoes in prior studies (30,31). As regards to the questionnaire, the high frequency of mosquito bites reported by the vendors reflects the high number of collected mosquitoes. The questionnaire also indicated a high level of mosquito-borne disease awareness, but a rather low perception of what diseases are spread by mosquitoes, only mentioning malaria to the highest degree, followed by dengue fever. This data contrasts with previous studies, showing 97% (Vientiane, 2013) (32) and 94% (Vientiane, 2011) (33) awareness of dengue fever. The Lao government has conducted educational campaigns to increase dengue fever knowledge, despite that, the knowledge level of this study remains low (34). Notably, none of the vendors mentioned JEV as a mosquito-borne disease.

In conclusion, the significant number of *Culex* adults and larvae found at the larger-sized markets underscores the role of stagnant wastewater as a significant factor contributing to mosquito breeding and prevalence. Despite no pathogenic flaviviruses were found, the conditions of LWMs create a high-risk environment for mosquito-borne disease emergence. Therefore, effective control measures, such as wastewater management, awareness campaigns, and vector control, are recommended to be implemented to mitigate the risks of mosquito-borne diseases associated with LWMs.

## Data Availability

The original data are provided in S2 table.

## Author contributions

Conceptualization, J.F. Lindahl and J. Ling; methodology, K. Vongphayloth, V. Phuttana, J.F. Lindahl, and J. Ling; validation, E. Asp, P. Höller, and J. Pärssinen; formal analysis, E. Asp, P. Höller, J. Pärssinen, N. Adsanycnah, and S. Chonephetsarath; investigation, E. Asp, P. Höller, and J. Pärssinen; resources, K. Vongphayloth, and V. Phuttana; data curation, E. Asp, J.F. Lindahl and J. Ling; writing—original draft preparation, E. Asp, J.F. Lindahl and J. Ling; writing—review and editing, all authors; visualization, E. Asp and J. Ling; supervision, J.F. Lindahl, and J. Ling; funding acquisition, M.M. Naguib, J.F. Lindahl, and J. Ling. All authors have read and agreed to the published version of the manuscript.

## Acknowledgements

This study was supported by FORMAS [2021–00833]. ‘Live and wet markets in Asia: the importance for food security and disease emergence’; J Ling is supported by the Swedish Research Council under Grant [2022–03219]. We thank the staff at the Faculty of Agriculture, National University of Laos for helping the translation of questionaries and the conduction of survey with vendors at markets, as well as Janina Krambrich for assisting with the qPCR, gel electrophoresis, and sequencing.

## Supporting Information Captions

S1 Table. Primers used for flavivirus typing and Japanese Encephalitis virus validation

JEV: Japanese Encephalitis virus. USUV: Usutu virus

S2 Table. Pooled adult mosquitoes and larvae/pupae mosquitoes

Overview of the pooled samples. Site_ID refers to a unique identifier for each sampling location. Pool 75, 83, 89, 108, 117, and 139 were blood-fed mosquitoes. Multiple sampling occurrences at the same site are indicated by appending a colon and an occurrence number.

S1 Fig. PCR products on gel electrophoresis

PCR products on gel electrophoresis. A representative figure from four of the gels made. The positive control (PC) of Usutu virus shows a band size of approximately 266 bp. Pools with bands of that size are considered a possible positive for flavivirus. The positive control (PC) of Usutu virus shows a band size of approximately 266 bp. Pools with bands of that size are considered a possible positive for flavivirus. The pool number is indicated first, followed by the sex and species of the pool. F.: female, M.: male, L.: larvae, Cx.qq.: Culex. quinquefasciaus, An.: Anopheles, NC.: Negative control.

S3 Table. Results from BLAST analysis of Sanger sequencing data

